# Medullary Cast Formation Predicts Kidney Outcomes Beyond Cortical Pathology

**DOI:** 10.64898/2026.03.10.26348108

**Authors:** Naruhiko Uchida, Kenji Tsuji, Hiroyuki Nakanoh, Kazuhiko Fukushima, Haruhito A. Uchida, Shinji Kitamura, Jun Wada

## Abstract

**Background:** Renal biopsy provides important prognostic information for patients with chronic kidney disease (CKD), primarily through evaluation of cortical histopathological lesions, including interstitial fibrosis and tubular atrophy (IFTA). However, the prognostic significance of renal medullary lesions remains poorly understood. We investigated whether medullary pathological findings are independently associated with renal outcomes and whether they improve prognostic discrimination beyond conventional cortical assessments.

**Methods:** This single-center retrospective cohort study screened 1,136 adult patients who underwent native kidney biopsy between 2011 and 2023. After applying predefined inclusion and exclusion criteria, 488 patients with adequate medullary tissue were included in the final analysis. Medullary fibrosis, inflammatory cell infiltration, and cast formation were semi-quantitatively graded and evaluated as predictors of renal outcomes. The primary outcome was a composite of ≥40% decline in estimated glomerular filtration rate (eGFR) or initiation of renal replacement therapy. Associations were assessed using Cox proportional hazards models with sequential adjustment for demographic factors, baseline eGFR, proteinuria, and cortical IFTA. Model discrimination was evaluated using Harrell’s concordance-index (C-index).

**Results:** During a median follow-up of 2.3 years, 112 patients (23.0%) reached the composite renal outcome. In multivariable analysis adjusted for age, sex, baseline eGFR, and proteinuria, medullary cast formation was significantly associated with adverse renal outcomes (hazard ratio [HR] 1.70, 95% confidence interval [CI] 1.28–2.24). This association remained significant after additional adjustment for IFTA (HR 1.64, 95% CI 1.21–2.21), whereas medullary fibrosis lost significance after IFTA adjustment. Addition of medullary cast formation improved C-index by 0.016, indicating incremental prognostic value beyond conventional predictors.

**Conclusion:** Medullary cast formation is independently associated with renal outcomes and improves prognostic discrimination beyond established cortical parameters. Systematic evaluation of medullary lesions during routine kidney biopsy may enhance risk stratification in CKD. Prospective studies are warranted to clarify the causal role of medullary pathology in CKD progression.

## Introduction

Renal biopsy remains the gold standard for the diagnosis and prognostic assessment of kidney diseases. While routine clinical parameters such as serum creatinine and proteinuria are essential for disease monitoring, histopathological findings provide critical information regarding structural damage, disease chronicity, and long-term renal prognosis^1–3^. Numerous studies have demonstrated that pathological evaluation refines risk stratification beyond clinical markers. For example, in IgA nephropathy, mesangial hypercellularity, endocapillary proliferation, segmental sclerosis, and tubular atrophy/interstitial fibrosis form the basis of the Oxford classification and are strongly associated with renal outcomes^4,5^. In addition, interstitial fibrosis and tubular atrophy (IFTA) have been consistently identified as robust predictors of renal function decline across various chronic kidney diseases (CKD)^6,7^. These observations underscore the central role of chronic tubulointerstitial damage in determining long-term prognosis. Moreover, certain drug-induced injuries preferentially affect the medullary region. For example, calcineurin inhibitors have been reported to induce fibrosis in the medullary rays, which may subsequently contribute to the progression of cortical IFTA^8^.

Importantly, conventional pathological assessment primarily focuses on cortical structures. In contrast, the renal medulla has been relatively understudied. The medulla, composed predominantly of loops of Henle, distal tubules, collecting ducts, and interstitium, is physiologically vulnerable to hypoxic injury, owing to its unique microvascular architecture and relatively low oxygen tension^9–12^. Previous experimental studies have highlighted the contribution of medullary hypoxia to CKD progression^13,14^, suggesting that medullary injury may reflect hemodynamic stress and ischemic burden not fully captured by cortical evaluation alone.

Histological changes in the medulla, including interstitial fibrosis, inflammatory cell infiltration, and intratubular cast formation, are frequently observed in routine biopsy specimens. Experimental models of obstructive nephropathy have demonstrated that tubular obstruction increases intratubular pressure, reduces peritubular capillary perfusion, activates inflammatory pathways, stimulates the renin-angiotensin-aldosterone (RAS) system, and ultimately promotes fibrosis^15,16^. Intratubular casts, particularly hyaline casts composed mainly of Tamm-Horsfall protein^17^, also known as uromodulin, may therefore represent not merely secondary findings but potentially active contributors to progressive tubular injury. Despite these mechanistic insights, the clinical prognostic significance of medullary histopathological findings has not been systematically investigated in heterogeneous CKD populations. It remains unclear whether medullary lesions provide independent prognostic information beyond established predictors such as eGFR, proteinuria, and cortical IFTA, or whether they simply reflect advanced cortical damage. We therefore hypothesized that medullary pathological findings are independently associated with renal outcomes. In addition, we examined whether incorporating medullary parameters improves prognostic discrimination beyond conventional cortical assessments.

## Methods

### Study Design and Participants

We conducted a single-center retrospective cohort study at Okayama University Hospital, Japan. A total of 1,136 patients underwent native renal biopsy between January 2011 and December 2023. Of these, 488 patients were included in the final analysis after excluding those younger than 18 years of age, those with an estimated glomerular filtration rate (eGFR) <15 mL/min/1.73 m² at the time of biopsy, and those with insufficient medullary tissue for pathological evaluation (Figure 1). The study protocol was approved by the institutional review board of Okayama University (Approval Nos. 1908-022 and 2407-038). The requirement for written informed consent was waived because of the retrospective design. Instead, information about the study was publicly disclosed on the institutional website, and patients were provided the opportunity to opt out.

**Figure 1.**
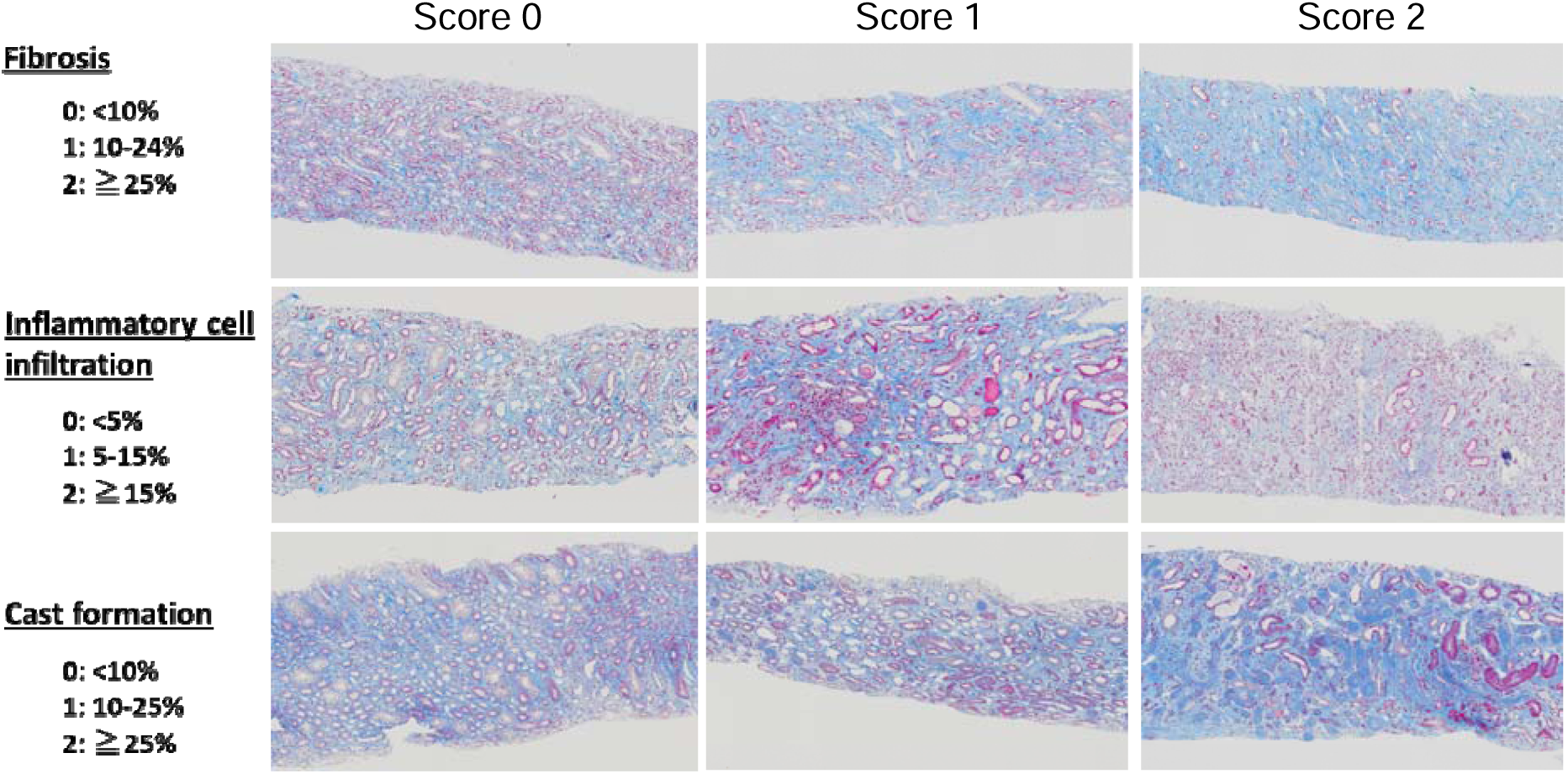
Medullary pathological findings and scoring system. Representative light microscopic images of renal medullary tissue stained with Masson’s trichrome showing medullary fibrosis, inflammatory cell infiltration, and cast formation. Each lesion was semi-quantitatively graded as 0, 1, or 2 based on severity.

### Histological Parameters

Renal tissue samples were obtained using 16– or 18-gauge biopsy needles and evaluated by light microscopy, immunofluorescence, and transmission electron microscopy. For light microscopic examination, all specimens were stained with Masson’s trichrome, periodic acid-Schiff (PAS), and periodic acid-methenamine silver (PAM). In the renal cortex, IFTA was semi-quantitatively graded according to the proportion of the cortical area involved as follows: 0 (<10%), 1 (10–24%), 2 (25–49%), and 3 (≥50%) according to previous reports and the Banff classification of kidney allograft pathology^6,18^. Vascular sclerosis, including interlobular arteries and arterioles, was graded based on the extent of involvement of the vessel wall circumference as follows: 0 (none), 1 (<25%), 2 (25–50%), and 3 (>50%). All biopsy specimens were independently reviewed at pathology conferences by multiple experienced nephrologists who were blinded to clinical outcomes, and discrepancies were resolved by consensus.

### Evaluation of Renal Medulla

Medullary pathological findings were evaluated using Masson’s trichrome staining, which allows clearer differentiation of fibrosis, inflammatory cell infiltration, and intratubular cast formation compared with other staining methods. Medullary lesions were semi-quantitatively assessed according to the proportion of the total medullary area involved in each biopsy specimen. Medullary fibrosis and cast formation were scored as 0 (<10%), 1 (10–24%), or 2 (≥25%), and inflammatory cell infiltration was scored as 0 (<5%), 1 (5–14%), or 2 (≥15%) (Figure 1). These criteria were adapted from the Banff classification of kidney allograft pathology^6,18^.

### Clinical Parameters

The following data at the time of renal biopsy were retrospectively collected from the electronic medical records of Okayama University Hospital: age, sex, body mass index (BMI), systolic and diastolic blood pressure, hemoglobin A1c (HbA1c), serum creatinine, eGFR calculated using the equation developed by the Japanese Society of Nephrology)^19^, proteinuria (urinary protein-to-creatinine ratio), hematuria (defined as ≥5 red blood cells per high-power field), smoking history (current/former/never), histopathological diagnosis, and baseline concomitant medications. These medications included RAS inhibitors (angiotensin receptor blockers (ARBs) and angiotensin-converting enzyme inhibitors (ACEIs)), calcium channel blockers (CCBs), and statins. HbA1c data are presented as National Glycohemoglobin Standardization Program values according to the recommendations of the Japanese Diabetes Society and the International Federation of Clinical Chemistry^20^.

### Predictors and Outcomes

The primary predictors were medullary pathological findings, including fibrosis, inflammatory cell infiltration, and cast formation, each graded on a scale of 0–2 as described above), as well as known predictive measurements, including IFTA scores^6^. The index date was defined as the date of renal biopsy. The study endpoint was a composite of (i) a ≥40% decline in eGFR from baseline and/or (ii) initiation of renal replacement therapy (RRT: dialysis or kidney transplantation)^21^. Participants were followed from the index date until the first occurrence of the endpoint or censored at the date of the last clinical visit.

### Statistical Analysis

Continuous variables were presented as mean ± standard deviation or median [interquartile range], and categorical variables as number (%). The cumulative incidence of renal events was estimated using the Kaplan-Meier method and compared using the log-rank test. Associations between medullary lesions and renal outcomes were evaluated using Cox proportional hazards models. Medullary scores (0–2) were treated as ordinal variables. Sequential adjustment was performed as follows: Model 0 (unadjusted), Model 1 (age and sex), Model 2 (Model 1 + eGFR and proteinuria), and Model 3 (Model 2 + IFTA). The proportional hazards assumption was evaluated by inspection of log-minus-log survival plots. Discriminative ability was assessed by Harrell’s concordance index (C-index)^22^. Associations between clinical and pathological parameters were examined using Spearman’s rank correlation coefficients, and independent associations were evaluated using ordinal logistic regression analysis. Analyses were performed with JMP (Student Edition version 18), with auxiliary computations for the C-index and spline modeling conducted in EZR. A two-sided p value < 0.05 was considered statistically significant.

## Results

### Clinical and Pathological Characteristics

A total of 488 patients were included in the final analysis (Figure 2). Baseline clinical characteristics and clinicopathological diagnoses of the study cohort are presented in Tables 1 and Supplemental Table 1-4. The mean age was 54.1 ± 17.9 years, and 217 patients (44.5%) were male. Hypertension was present in 227 patients (46.5%), and diabetes mellitus in 121 patients (24.8%). The mean serum creatinine level was 1.06 ± 0.50 mg/dL, and the mean eGFR was 60.9 ± 26.3 mL/min/1.73 m². The median urinary protein excretion was 1.1 g/gCr (interquartile range [IQR], 0.4–3.1). The most common diagnosis was IgA nephropathy (31.5 %), followed by ANCA-associated vasculitis (7.6 %) and lupus nephritis (7.2 %); detailed diagnoses are shown in Supplemental Table 5. Medullary pathological lesions were frequently observed in biopsy specimens, including fibrosis, inflammatory cell infiltration, and intratubular cast formation with varying degrees of severity (Figure 2 and Table 3).

**Figure 2.**
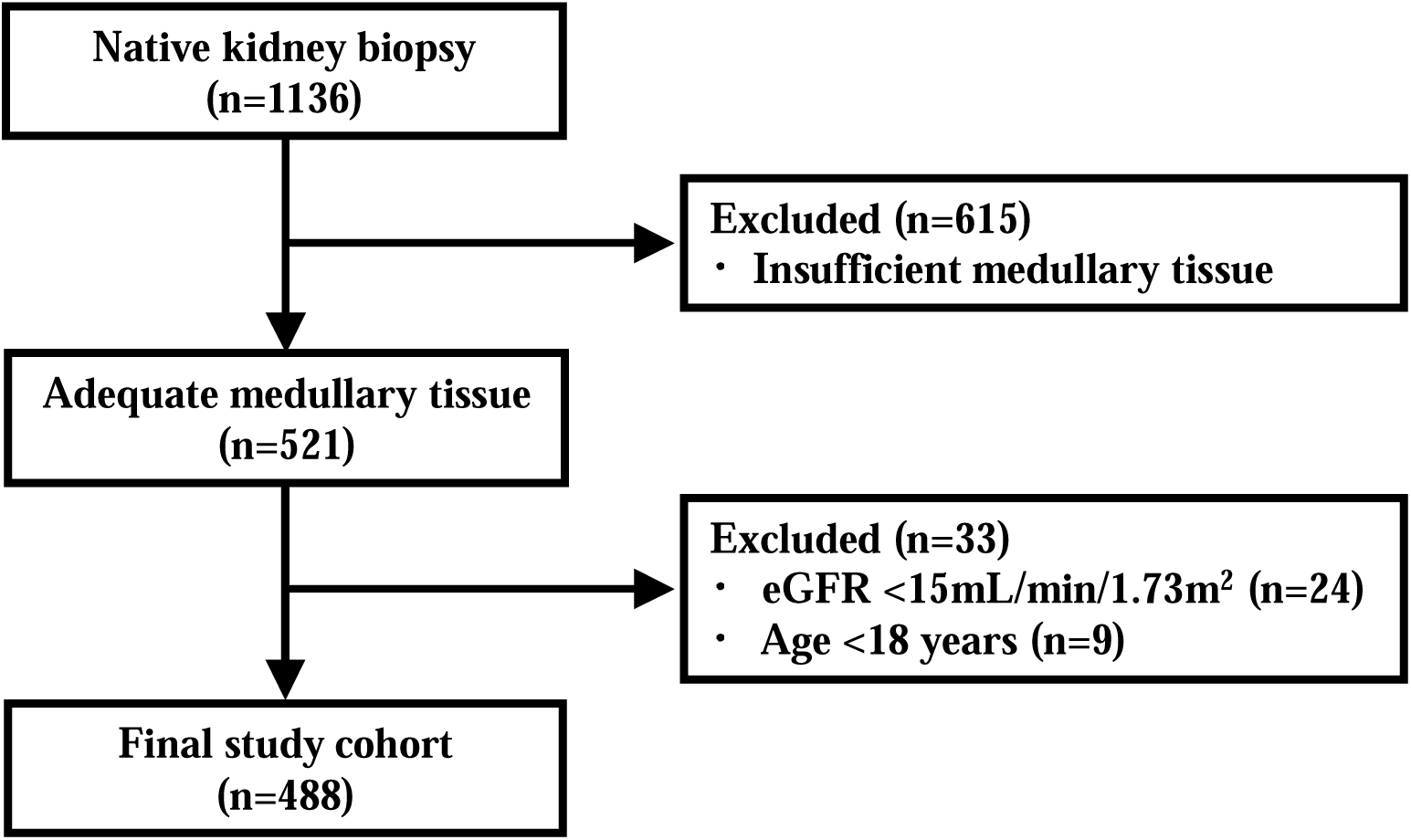
Flowchart of study participants. eGFR, estimated glomerular filtration rate.

### Prognostic Value of Medullary Pathological Findings

During a median follow-up period of 2.3 years, 112 patients experienced the composite renal outcome. Kaplan-Meier analyses demonstrated significant differences in renal outcomes according to the severity of medullary fibrosis, medullary cast formation, and cortical IFTA (Figure 3). Multivariable Cox proportional hazards models were used to evaluate the independent associations between medullary pathological findings and renal outcomes (Table 2). In Model 2, adjusted for age, sex, baseline eGFR, and proteinuria, both medullary fibrosis (hazard ratio [HR], 1.32; 95% confidence interval [CI], 1.01–1.71) and medullary cast formation (HR, 1.70; 95% CI, 1.28–2.24) were significantly associated with renal outcomes. After additional adjustment for cortical IFTA (Model 3), medullary cast formation remained significantly associated with renal outcomes (HR, 1.64; 95% CI, 1.21–2.21), whereas the association between medullary fibrosis and renal outcomes was attenuated. These findings indicate that medullary cast formation provides prognostic information beyond established cortical chronic damage. In contrast, medullary inflammatory cell infiltration was not significantly associated with renal outcomes even in Model 1 (adjusted for age and sex; HR, 3.27; 95% CI, 0.79–12.10). Multicollinearity among covariates was assessed using variance inflation factors, and no substantial multicollinearity was observed (all VIFs < 2.5).

**Figure 3.**
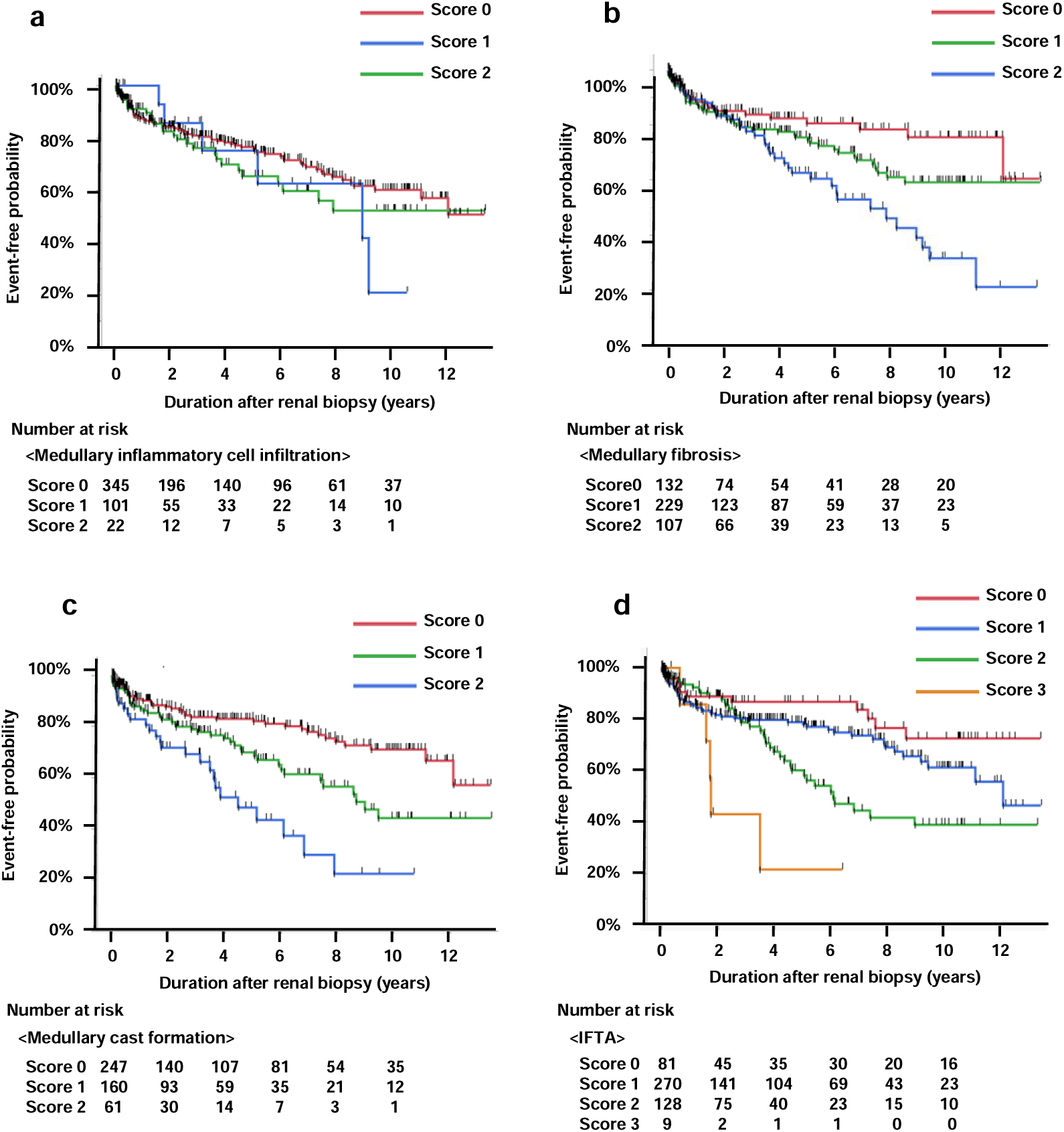
Renal survival rate stratified by medullary findings and IFTA score. (a) Renal survival rate stratified by medullary inflammatory cell infiltration. Log rank test: *p* = 0.45. (b) Renal survival rate stratified by medullary fibrosis. Log rank test: *p* = 0.002. (c) Renal survival rate stratified by medullary cast formation. Log rank test: *p* <0.001. (d) Renal survival rate stratified by IFTA score. Log rank test: *p* <0.001. IFTA, interstitial fibrosis and tubular atrophy.

**Table 1.** Clinical and histopathological characteristics for all patients.

| Characteristics | n=488 |  |
| --- | --- | --- |
| Men | 218 (44.5%) |  |
| Age, years | 54.1±17.9 |  |
| BMI, kg/m<sup>2</sup> | 22.7±4.0 |  |
| sBP, mmHg | 127.4±18.7 |  |
| dBp, mmHg | 79.4±18.7 |  |
| sCr, mg/dL | 1.06±0.50 |  |
| eGFR, ml/min/1.73 m<sup>2</sup> | 60.9±26.3 |  |
| HbA1c, % | 5.7 (5.4-6.1) |  |
| U-P, g/gCr | 1.14 (0.38-3.11) |  |
| Hematuria, % | 289 (59.2%) |  |
| Urine cast | 266 (46.3%) |  |
| Hypertension | 227 (46.5%) |  |
| DM | 121 (24.8%) |  |
| ACE-I or ARB | 171 (35.0%) |  |
| Ca blocker | 169 (34.6%) |  |
| Statins | 116 (23.8%) |  |
| Smoking | 211 (43.2%) |  |
| IFTA | 0 | 81 (16.6%) |
|  | 1 | 270 (55.3%) |
|  | 2 | 128 (26.2%) |
|  | 3 | 9 (1.8%) |
| Medullary fibrosis | 0 | 135 (27.7%) |
|  | 1 | 239 (49.0%) |
|  | 2 | 114 (23.4%) |
| Medullary inflammatory cell infiltration | 0 | 359 (73.6%) |
|  | 1 | 107 (21.9%) |
|  | 2 | 22 (4.5%) |
| Medullary cast formation | 0 | 254 (52.0%) |
|  | 1 | 170 (34.8%) |
|  | 2 | 64 (13.1%) |

Values reported as n (%) for categorical variables and mean ±SD or median (IQR) for continuous variables. BMI, body mass index; sBP, systolic blood pressure; dBp, diastolic blood pressure; sCr, serum creatinine; eGFR, estimated glomerular filtration rate; HbA1c, hemoglobin A1c; U-P, urinary protein excretion; DM, diabetes mellitus; ACE-I, angiotensin-converting enzyme inhibitor; ARB, angiotensin II type 1 receptor blocker; IFTA, interstitial fibrosis and tubular atrophy.

**Table 2.** Cox proportional hazards analysis of renal medullary pathological findings and IFTA.

|  | Model 0<br>HR (95%CI) | Model 1<br>HR (95%CI) | Model 2<br>HR (95%CI) | Model 3<br>HR (95%CI) |
| --- | --- | --- | --- | --- |
| Medullary fibrosis | 1.58<br>(1.22-2.05) | 1.44<br>(1.09-1.90) | 1.38<br>(1.02-1.87) | 1.21<br>(0.87-1.68) |
| Medullary inflammatory cell infiltration | 4.45<br>(1.15-15.46) | 3.27<br>(0.79-12.10) | 2.13<br>(0.40-10.16) | 0.31<br>(0.03-2.53) |
| Medullary cast formation | 1.86<br>(1.45-2.38) | 1.75<br>(1.35-2.27) | 1.70<br>(1.28-2.24) | 1.56<br>(1.16-2.10) |
| IFTA | 1.68<br>(1.28-2.19) | 1.57<br>(1.18-2.09) | 1.61<br>(1.15-2.27) |  |

Model 0: No adjustment factors. Model 1: Sex, Age. Model 2: Model 1 + eGFR, Urinary protein. Model 3: Model 2 + IFTA. HR, hazard ratio; 95%CI, 95% confidence interval; IFTA, interstitial fibrosis and tubular atrophy.

To evaluate whether medullary pathological findings improved prognostic discrimination beyond conventional risk factors, C-index was calculated. A model including age, sex, eGFR, and proteinuria yielded a C-index of 0.634 (standard error [SE], 0.030). Incorporating cortical IFTA increased the C-index to 0.643 (SE, 0.028). Further inclusion of medullary cast formation increased the C-index to 0.655 (SE, 0.028), indicating incremental improvement in prognostic discrimination.

In a subgroup analysis restricted to patients with IgA nephropathy (n = 154), 28 renal outcome events occurred during a median follow-up of 3.4 years. Kaplan-Meier analyses demonstrated significant differences in renal outcomes according to the severity of medullary fibrosis and cast formation (Supplemental Figure 1). These findings were consistent with the results observed in the overall cohort. Multivariable Cox analyses were not performed because of the limited number of outcome events in this subgroup.

### Associations Between Medullary Pathological Findings and Clinical / Pathological Parameters

Associations between medullary lesions and established clinical and pathological prognostic factors were examined using Spearman’s rank correlation analysis (Supplemental Table 6). Medullary fibrosis showed a positive correlation with medullary cast formation (ρ = 0.54, p < 0.001) and cortical IFTA (ρ = 0.56, p < 0.001), and a negative correlation with eGFR (ρ = −0.53, p < 0.001). Similarly, medullary cast formation was positively correlated with cortical IFTA (ρ = 0.49, p < 0.001) and negatively correlated with eGFR (ρ = −0.48, p < 0.001). Urinary protein excretion showed only weak correlations with medullary fibrosis (ρ = 0.11, p = 0.017) and medullary cast formation (ρ = 0.14, p = 0.002).

To further evaluate independent relationships, ordinal logistic regression analyses were performed (Table 3). Medullary fibrosis was independently associated with age (odds ratio [OR], 1.03; 95% CI, 1.01–1.05), medullary cast formation (OR, 3.06; 95% CI, 2.16–3.98), and cortical IFTA (OR, 2.70; 95% CI, 1.85–3.98). Medullary cast formation was independently associated with medullary fibrosis (OR, 3.31; 95% CI, 2.27–4.86) and cortical IFTA (OR, 2.27; 95% CI, 1.55–3.36), and inversely associated with eGFR (OR, 0.81; 95% CI, 0.72–0.92 per 10 mL/min/1.73 m² increase). Consistently, cortical IFTA was positively associated with both medullary fibrosis (OR, 2.72; 95% CI, 1.85–4.03) and medullary cast formation (OR, 2.03; 95% CI, 1.44–2.88). Taken together, these findings indicate that although medullary lesions correlate with established markers of chronic kidney damage, medullary cast formation retains independent prognostic significance beyond cortical IFTA.

**Table 3.** Ordinal logistic regression analysis of medullary pathological findings and established prognostic factors.

|  | Medullary fibrosis |  | Medullary cast formation |  | IFTA |  |
| --- | --- | --- | --- | --- | --- | --- |
|  | Estimated Odds (95%CI) | p-value | Estimated Odds (95%CI) | p-value | Estimated Odds (95%CI) | p-value |
| Age | 1.03<br>(1.01-1.05) | <0.001 | 1.00<br>(0.99-1.02) | 0.54 | 0.99<br>(0.98-1.01) | 0.21 |
| eGFR (10 increase) | 0.93<br>(0.83-1.04) | 0.18 | 0.81<br>(0.72-0.92) | <0.001 | 0.70<br>(0.62-0.78) | <0.001 |
| U-P | 1.02<br>(0.96-1.09) | 0.50 | 1.03<br>(0.96-1.09) | 0.40 | 0.99<br>(0.93-1.05) | 0.70 |
| Medullary fibrosis |  |  | 3.31<br>(2.27-4.86) | <0.001 | 2.72<br>(1.85-4.03) | <0.001 |
| Medullary cast formation | 3.06<br>(2.16-4.37) | <0.001 |  |  | 2.03<br>(1.44-2.88) | <0.001 |
| IFTA | 2.70<br>(1.85-3.98) | <0.001 | 2.27<br>(1.55-3.36) | <0.001 |  |  |

95%CI, 95% confidence interval; eGFR, estimated glomerular filtration rate; U-P, urinary protein excretion; IFTA, interstitial fibrosis and tubular atrophy.

## Discussion

In this retrospective cohort study of patients undergoing kidney biopsy, we demonstrated that medullary cast formation was independently associated with an increased risk of adverse renal outcomes after adjustment for age, sex, baseline eGFR, proteinuria, and cortical IFTA. In addition, incorporating medullary cast formation into prognostic models resulted in incremental improvement in the C-index, suggesting additional prognostic value beyond established clinical and pathological predictors. These findings indicate that renal medullary pathology may provide prognostic information beyond conventional cortical evaluation.

Previous studies have consistently shown that cortical chronic damage, particularly IFTA, is strongly associated with renal prognosis across various kidney diseases^6,7^. In contrast, medullary lesions have rarely been quantitatively incorporated into prognostic analyses in clinical cohorts. Experimental studies have suggested that medullary hypoxia and tubular obstruction play important roles in CKD progression^13,15^, but clinical evidence linking medullary histopathology to long-term renal outcomes has remained limited. The present findings help bridge this gap by demonstrating a significant association between medullary cast formation and renal function decline in a real-world kidney biopsy cohort.

The renal medulla is particularly vulnerable to injury because of its relatively low oxygen tension and high metabolic demand^12^. Tubular obstruction within this region may therefore have amplified consequences. Intratubular cast formation can increase intraluminal pressure, impair tubular flow, and reduce peritubular capillary perfusion. Experimental models of obstructive nephropathy have demonstrated that sustained elevation of intratubular pressure induces tubular epithelial injury, interstitial inflammation, and progressive fibrosis through activation of hypoxia-inducible pathways and the RAS system^15^. These observations suggest that intratubular casts may act as a form of partial tubular obstruction, analogous to mechanisms observed in experimental models of obstructive nephropathy (Figure 4). Most casts observed in this study were predominantly hyaline casts, primarily composed of uromodulin produced in the thick ascending limb. Uromodulin polymerization is influenced by urinary concentration, ionic strength, and pH, aggregating more readily under acidic conditions^17,23,24^. Thus, medullary cast formation may reflect not only protein overload but also alterations in the tubular microenvironment and urinary chemistry.

**Figure 4.**
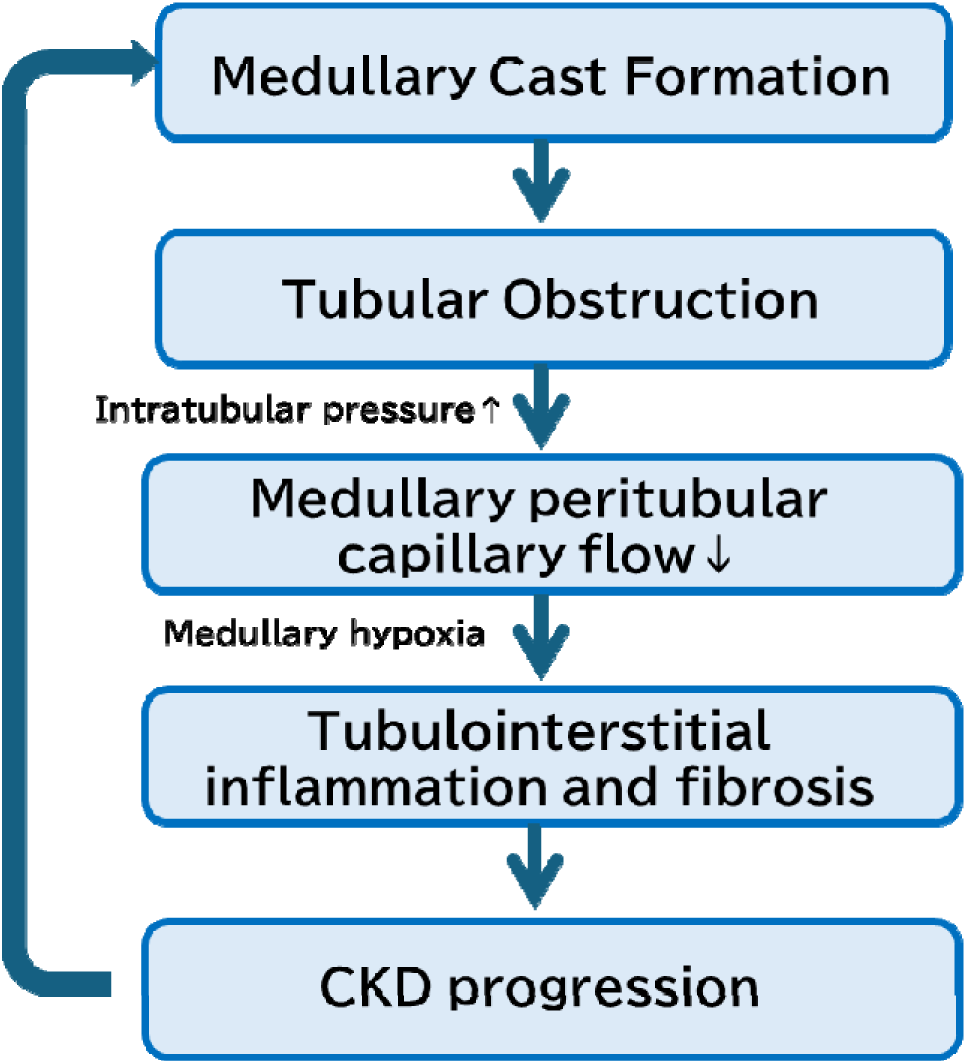
Proposed mechanism linking medullary cast formation to CKD progression. Medullary cast formation may promote tubular obstruction, leading to increased intratubular pressure and reduced medullary peritubular capillary flow. These changes may induce medullary hypoxia and subsequent tubulointerstitial inflammation and fibrosis, thereby contributing to CKD progression. Cast formation may also be facilitated by alterations in urinary concentration and chemistry within the renal medulla. This conceptual model is based on the findings of the present study and prior experimental evidence. CKD, chronic kidney disease.

Importantly, medullary cast formation remained significantly associated with renal outcomes even after adjustment for cortical IFTA, whereas the association between medullary fibrosis and renal outcomes was attenuated. This observation suggests that cast formation may represent a dynamic pathological process distinct from static chronic fibrosis rather than merely reflecting advanced cortical damage. The observed negative association between cast formation and eGFR raises the possibility of a bidirectional relationship, in which declining renal function promotes cast formation, which in turn exacerbates tubular injury and accelerates renal deterioration. From a practical standpoint, medullary tissue is frequently obtained incidentally during routine kidney biopsy. However, it is rarely incorporated into structured prognostic evaluation. The present findings suggest that systematic assessment of medullary cast formation may enhance risk stratification without requiring additional invasive procedures. This may be particularly useful when cortical sampling is limited, but even in adequately sampled biopsies, medullary parameters provided additive prognostic information.

Several limitations should be acknowledged. First, this was a single-center retrospective study with a relatively limited sample size, and external validation is required. Second, medullary lesions were assessed using semi-quantitative scoring, which may introduce observer-dependent variability. Evaluation of medullary fibrosis is particularly challenging because the medullary interstitium physiologically contains a relatively higher baseline proportion of fibrous components compared with the cortex, making distinction between normal structural composition and pathological fibrosis difficult. Third, sampling bias represents an important concern. Not all biopsy specimens contain sufficient medullary tissue, and the anatomical heterogeneity of the medulla, including the outer stripe and inner stripe of the outer medulla as well as the inner medulla, may influence pathological findings. In addition, most medullary tissue obtained by kidney biopsy likely corresponds to the outer medulla adjacent to the cortex, because biopsy procedures primarily target cortical structures. Differences in sampling location could therefore affect lesion severity and limit uniform assessment. Fourth, the study cohort included heterogeneous kidney diseases. Although this enhances generalizability, disease-specific mechanisms and treatment responses may influence the observed associations. Larger multicenter studies enabling disease-specific subgroup analyses will be necessary to clarify whether the prognostic significance of medullary lesions differs across underlying etiologies. Finally, as an observational study, causal relationships between medullary cast formation and renal decline cannot be definitively established.

In conclusion, medullary cast formation was independently associated with an increased risk of adverse renal outcomes and improved prognostic discrimination beyond established cortical parameters. Incorporating structured evaluation of medullary pathology into routine kidney biopsy assessment may enhance prognostic stratification in CKD. Prospective and mechanistic studies are warranted to determine whether medullary lesions represent potential therapeutic targets in the progression of CKD.

### Disclosures

All the authors declared no competing interests.

## Funding

This work was supported by the Japanese Society for the Promotion of Science (JSPS)/Grant-in-Aid for Scientific Research (C) (grant no. 24K11411). The funder has no influence on the design or analyses in this study.

## Data Availability

All data produced in the present study are available from the corresponding author upon reasonable request.

## Acknowledgements

The authors used ChatGPT (OpenAI, San Francisco, CA, USA) for English language editing and manuscript refinement. The authors take full responsibility for the content of this manuscript.

## Author contributions

Conceptualization, K.T.; formal analysis, K.T., H.N., N.U. and K.F.; investigation, K.T., H.N. and K.F.; writing—original draft preparation, K.T.; writing—review and editing, K.T., H.N., K.F., HA. U., S.K. and J.W.; supervision, J.W.; All authors have read and agreed to the published version of the manuscript.

## Data Sharing Statement

The data that support the findings of this study are available from the corresponding author upon reasonable request.

## Supplemental Material

### Supplemental File

**Supplemental Table 1.** Baseline characteristics according to medullary inflammatory cell infiltration

**Supplemental Table 2.** Baseline characteristics according to medullary fibrosis

**Supplemental Table 3.** Baseline characteristics according to medullary cast formation

**Supplemental Table 4.** Baseline characteristics according to IFTA

**Supplemental Table 5.** Clinicopathologic diagnoses of the study population

**Supplemental Table 6.** Spearman’s correlation coefficients among clinical and pathological variables

**Supplemental Figure 1**. Renal survival rate in patients with IgA nephropathy

## Notes

### Competing Interest Statement

The authors have declared no competing interest.

### Author Declarations

The study protocol was approved by the institutional review board of Okayama University (Approval Nos. 1908-022 and 2407-038). The requirement for written informed consent was waived because of the retrospective design.

