## Supplemental Materials for "Medullary Cast Formation Predicts Kidney Outcomes Beyond Cortical Pathology"

### Supplemental Material

#### Table of contents

**1. Supplemental Table**

**Supplemental Table 1.** Baseline characteristics according to medullary inflammatory cell infiltration

**Supplemental Table 2.** Baseline characteristics according to medullary fibrosis

**Supplemental Table 3.** Baseline characteristics according to medullary cast formation

**Supplemental Table 4.** Baseline characteristics according to IFTA

**Supplemental Table 5.** Clinicopathologic diagnoses of the study population

**Supplemental Table 6.** Spearman's correlation coefficients among clinical and pathological variables

**2. Supplemental Figure**

**Supplemental Figure 1.** Renal survival rate in patients with IgA nephropathy

**3. STROBE Statement**

**Supplemental Table 1. Baseline characteristics according to medullary inflammatory cell infiltration**

| | | All patients (n=488) | Medullary inflammatory cell infiltration | | | p for trend |
|---|---|---|---|---|---|---|
| | | | 0 (n=359) | 1 (n=107) | 2 (n=22) | |
| Men | | 218 (44.5%) | 175 (48.7%) | 36 (33.6%) | 15 (31.8%) | 0.003 |
| Age, years | | 54.1±17.9 | 52.8±18.3 | 58.0±16.2 | 56.6±16.5 | 0.011 |
| BMI, kg/$m^2$ | | 22.7±4.0 | 22.7±4.1 | 22.7±3.8 | 22.6±3.5 | 0.83 |
| sBP, mmHg | | 127.4±18.7 | 126.0±18.5 | 131.1±18.2 | 131.2±21.8 | 0.005 |
| dBP, mmHg | | 79.4±18.7 | 79.4±12.1 | 79.5±12.3 | 79.4±19.8 | 0.91 |
| sCr, mg/dL | | 1.06±0.50 | 0.96±0.39 | 1.31±0.64 | 1.55±0.50 | <0.001 |
| eGFR, mL/min/1.73 $m^2$ | | 60.9±26.3 | 66.5±25.6 | 47.5±22.6 | 34.1±12.4 | <0.001 |
| HbA1c, % | | 5.7 (5.4-6.1) | 5.7 (5.4-6.1) | 5.8 (5.5-6.1) | 5.8 (5.2-6.4) | 0.44 |
| U-P, g/gCr | | 1.14 (0.38-3.11) | 1.02 (0.35-3.09) | 1.36 (0.46-3.19) | 1.30 (0.79-2.70) | 0.13 |
| Hematuria, % | | 289 (59.2%) | 211 (58.8%) | 66 (61.7%) | 12 (54.5%) | 0.79 |
| Urine cast | | 266 (46.3%) | 164 (45.7%) | 51 (47.7%) | 11 (50.0%) | 0.63 |
| Hypertension | | 227 (46.5%) | 154 (42.9%) | 61 (57.0%) | 12 (54.5%) | 0.009 |
| DM | | 121 (24.8%) | 86 (24.0%) | 28 (26.2%) | 7 (31.8%) | 0.44 |
| ACE-I or ARB | | 171 (35.0%) | 116 (32.3%) | 45 (42.1%) | 10 (45.5%) | 0.033 |
| Ca blocker | | 169 (34.6%) | 115 (32.0%) | 42 (39.3%) | 12 (54.5%) | 0.031 |
| Statins | | 116 (23.8%) | 79 (22.0%) | 32 (29.9%) | 5 (22.7%) | 0.38 |
| Smoking | | 211 (43.2%) | 161 (44.8%) | 40 (37.4%) | 10 (45.5%) | 0.21 |
| IFTA | 0 | 81 (16.6%) | 78 (21.7%) | 3 (2.8%) | 0 (0.0%) | <0.001 |
| | 1 | 270 (55.3%) | 217 (60.4%) | 52 (48.6%) | 1 (4.5%) | |
| | 2 | 128 (26.2%) | 62 (17.3%) | 50 (46.7%) | 16 (72.7%) | |
| | 3 | 9 (1.8%) | 2 (0.5%) | 2 (1.9%) | 5 (22.7%) | |
| Medullary fibrosis | 0 | 135 (27.7%) | 133 (37.0%) | 2 (1.9%) | 0 (0.0%) | <0.001 |
| | 1 | 239 (49.0%) | 178 (49.6%) | 59 (55.1%) | 2 (9.1%) | |
| | 2 | 114 (23.4%) | 48 (13.4%) | 46 (43.0%) | 20 (90.9%) | |
| Medullary cast formation | 0 | 254 (52.0%) | 216 (60.2%) | 32 (29.9%) | 6 (27.3%) | <0.001 |
| | 1 | 170 (34.8%) | 113 (31.5%) | 49 (45.8%) | 8 (36.4%) | |
| | 2 | 64 (13.1%) | 30 (8.4%) | 26 (24.3%) | 8 (36.4%) | |

BMI, body mass index; sBP, systolic blood pressure; dBP, diastolic blood pressure; sCr, serum creatinine; eGFR, estimated glomerular filtration rate; HbA1c, hemoglobin A1c; U-P, urinary protein excretion; DM, diabetes mellitus; ACE-I, angiotensin–converting enzyme inhibitor; ARB, angiotensin II type 1 receptor blocker; IFTA, interstitial fibrosis and tubular atrophy.

**Supplemental Table 2. Baseline characteristics according to medullary fibrosis**

| | | All patients (n=488) | Medullary fibrosis | | | p for trend |
|---|---|---|---|---|---|---|
| | | | 0 (n=135) | 1 (n=239) | 2 (n=114) | |
| Men | | 218 (44.5%) | 60 (44.4%) | 109 (45.6%) | 49 (43.0%) | 0.84 |
| Age, years | | 54.1±17.9 | 41.7±16.8 | 57.5±16.3 | 61.8±14.6 | <0.001 |
| BMI, $kg/m^2$ | | 22.7±4.0 | 22.7±4.5 | 22.6±3.7 | 22.8±3.9 | 0.48 |
| sBP, mmHg | | 127.4±18.7 | 120.6±18.5 | 129.5±18.1 | 130.9±18.1 | <0.001 |
| dBP, mmHg | | 79.4±18.7 | 78.7±12.2 | 79.7±12.1 | 79.6±13.7 | 0.59 |
| sCr, mg/dL | | 1.06±0.50 | 0.80±0.26 | 1.06±0.47 | 1.38±0.57 | <0.001 |
| eGFR, mL/min/1.73 $m^2$ | | 60.9±26.3 | 81.0±26.4 | 58.2±22.6 | 42.6±18.7 | <0.001 |
| HbA1c, % | | 5.7 (5.4-6.1) | 5.6 (5.3-5.9) | 5.8 (5.5-6.2) | 5.8 (5.5-6.2) | <0.001 |
| U-P, g/gCr | | 1.14 (0.38-3.11) | 0.92 (0.31-2.19) | 1.06 (0.35-3.11) | 1.45 (0.51-3.69) | 0.016 |
| Hematuria, % | | 289 (59.2%) | 85 (63.0%) | 144 (60.3%) | 60 (52.6%) | 0.11 |
| Urine cast | | 266 (46.3%) | 58 (43.0%) | 117 (49.0%) | 51 (44.7%) | 0.71 |
| Hypertension | | 227 (46.5%) | 33 (24.4%) | 123 (51.5%) | 71 (62.3%) | <0.001 |
| DM | | 121 (24.8%) | 32 (23.7%) | 61 (25.5%) | 28 (24.6%) | 0.84 |
| ACE-I or ARB | | 171 (35.0%) | 29 (21.5%) | 89 (37.2%) | 53 (46.5%) | <0.001 |
| Ca blocker | | 169 (34.6%) | 24 (17.8%) | 87 (36.4%) | 58 (50.9%) | <0.001 |
| Statins | | 116 (23.8%) | 22 (16.3%) | 56 (23.4%) | 38 (33.3%) | 0.004 |
| Smoking | | 211 (43.2%) | 53 (39.3%) | 105 (43.9%) | 53 (46.5%) | 0.42 |
| IFTA | 0 | 81 (16.6%) | 61 (45.2%) | 18 (7.5%) | 2 (1.8%) | <0.001 |
| | 1 | 270 (55.3%) | 69 (51.1%) | 160 (66.9%) | 41 (36.0%) | |
| | 2 | 128 (26.2%) | 5 (3.7%) | 60 (25.1%) | 63 (55.3%) | |
| | 3 | 9 (1.8%) | 0 (0.0%) | 1 (0.4%) | 8 (7.0%) | |
| Medullary inflammatory cell infiltration | 0 | 359 (73.6%) | 133 (98.5%) | 178 (74.5%) | 48 (42.1%) | <0.001 |
| | 1 | 107 (21.9%) | 2 (1.5%) | 59 (24.7%) | 46 (40.4%) | |
| | 2 | 22 (4.5%) | 0 (0.0%) | 2 (0.8%) | 20 (17.5%) | |
| Medullary cast formation | 0 | 254 (52.0%) | 118 (87.4%) | 116 (48.5%) | 20 (17.5%) | <0.001 |
| | 1 | 170 (34.8%) | 16 (11.9%) | 103 (43.1%) | 51 (44.7%) | |
| | 2 | 64 (13.1%) | 1 (0.7%) | 20 (8.4%) | 43 (37.7%) | |

BMI, body mass index; sBP, systolic blood pressure; dBP, diastolic blood pressure; sCr, serum creatinine; eGFR, estimated glomerular filtration rate; HbA1c, hemoglobin A1c; U-P, urinary protein excretion; DM, diabetes mellitus; ACE-I, angiotensin–converting enzyme inhibitor; ARB, angiotensin II type 1 receptor blocker; IFTA, interstitial fibrosis and tubular atrophy.

**Supplemental Table 3. Baseline characteristics according to medullary cast formation**

| | | All patients (n=488) | Medullary cast formation | | | p for trend |
|---|---|---|---|---|---|---|
| | | | 0 (n=254) | 1 (n=170) | 2 (n=64) | |
| Men | | 218 (44.5%) | 111 (43.7%) | 77 (45.3%) | 30 (46.9%) | 0.62 |
| Age, years | | 54.1±17.9 | 49.0±18.1 | 58.4±15.9 | 63.2±15.1 | <0.001 |
| BMI, kg/m$^2$ | | 22.7±4.0 | 22.8±3.9 | 22.6±4.0 | 22.2±4.3 | 0.39 |
| sBP, mmHg | | 127.4±18.7 | 124.3±18.7 | 131.5±18.3 | 128.3±17.7 | 0.001 |
| dBP, mmHg | | 79.4±18.7 | 78.7±12.3 | 81.4±12.6 | 76.8±12.4 | 0.59 |
| sCr, mg/dL | | 1.06±0.50 | 0.89±0.40 | 1.18±0.46 | 1.44±0.61 | <0.001 |
| eGFR, mL/min/1.73 m$^2$ | | 60.9±26.3 | 72.5±26.2 | 50.8±20.0 | 41.6±18.3 | <0.001 |
| HbA1c, % | | 5.7 (5.4-6.1) | 5.7 (5.3-6.1) | 5.8 (5.4-6.2) | 5.8 (5.5-6.1) | 0.057 |
| U-P, g/gCr | | 1.14 (0.38-3.11) | 0.83 (0.28-2.78) | 1.28 (0.54-2.97) | 1.70 (0.53-3.83) | 0.002 |
| Hematuria, % | | 289 (59.2%) | 149 (58.7%) | 103 (60.6%) | 37 (57.8%) | 0.89 |
| Urine cast | | 266 (46.3%) | 101 (39.8%) | 90 (52.9%) | 35 (54.7%) | 0.003 |
| Hypertension | | 227 (46.5%) | 97 (38.2%) | 90 (52.9%) | 40 (62.5%) | <0.001 |
| DM | | 121 (24.8%) | 66 (26.0%) | 41 (24.1%) | 14 (21.9%) | 0.48 |
| ACE-I or ARB | | 171 (35.0%) | 69 (27.2%) | 71 (41.8%) | 31 (48.4%) | <0.001 |
| Ca blocker | | 169 (34.6%) | 54 (26.4%) | 69 (40.6%) | 33 (51.6%) | <0.001 |
| Statins | | 116 (23.8%) | 54 (21.3%) | 37 (21.8%) | 25 (39.1%) | 0.016 |
| Smoking | | 211 (43.2%) | 99 (39.0%) | 81 (47.6%) | 31 (48.4%) | 0.12 |
| IFTA | 0 | 81 (16.6%) | 74 (29.1%) | 7 (4.1%) | 0 (0.0%) | <0.001 |
| | 1 | 270 (55.3%) | 152 (59.8%) | 93 (54.7%) | 25 (39.0%) | |
| | 2 | 128 (26.2%) | 27 (10.6%) | 68 (40.0%) | 33 (51.6%) | |
| | 3 | 9 (1.8%) | 1 (0.4%) | 2 (1.2%) | 6 (9.4%) | |
| Medullary fibrosis | 0 | 135 (27.7%) | 118 (46.5%) | 16 (9.4%) | 1 (1.6%) | <0.001 |
| | 1 | 239 (49.0%) | 116 (45.7%) | 103 (60.6%) | 20 (31.3%) | |
| | 2 | 114 (23.4%) | 20 (7.9%) | 51 (30.0%) | 43 (67.2%) | |
| Medullary inflammatory cell infiltration | 0 | 359 (73.6%) | 216 (85.0%) | 113 (66.5%) | 30 (46.9%) | <0.001 |
| | 1 | 107 (21.9%) | 32 (12.6%) | 49 (28.8%) | 26 (40.6%) | |
| | 2 | 22 (4.5%) | 6 (2.4%) | 8 (4.7%) | 8 (12.5%) | |

BMI, body mass index; sBP, systolic blood pressure; dBP, diastolic blood pressure; sCr, serum creatinine; eGFR, estimated glomerular filtration rate; HbA1c, hemoglobin A1c; U-P, urinary protein excretion; DM, diabetes mellitus; ACE-I, angiotensin–converting enzyme inhibitor; ARB, angiotensin II type 1 receptor blocker; IFTA, interstitial fibrosis and tubular atrophy.

**Supplemental Table 4. Baseline characteristics according to IFTA**

| | | All patients (n=488) | IFTA | | | | p for trend |
|---|---|---|---|---|---|---|---|
| | | | 0 (n=81) | 1 (n=270) | 2 (n=128) | 3 (n=9) | |
| Men | | 218 (44.5%) | 36 (44.4%) | 126(46.7%) | 51 (39.8%) | 5 (55.6%) | 0.50 |
| Age, years | | 54.1±17.9 | 40.7±17.1 | 54.8±17.0 | 61.2±14.8 | 55.6±18.0 | <0.001 |
| BMI, kg/m$^2$ | | 22.7±4.0 | 22.8±4.2 | 22.5±3.66 | 22.5±4.49 | 23.5±3.92 | 0.79 |
| sBP, mmHg | | 127.4±18.7 | 119.5±15.5 | 127.1±19.0 | 132.2±18.2 | 135.7±17.9 | <0.001 |
| dBP, mmHg | | 79.4±18.7 | 77.2±10.5 | 79.8±12.1 | 80.4±14.2 | 75.9±15.6 | 0.23 |
| sCr, mg/dL | | 1.06±0.50 | 0.78±0.27 | 0.96±0.40 | 1.41±0.55 | 1.90±0.50 | <0.001 |
| eGFR, mL/min/1.73 m$^2$ | | 60.9±26.3 | 84.8±26.6 | 64.2±22.2 | 40.9±17.3 | 28.9±7.66 | <0.001 |
| HbA1c, % | | 5.7 (5.4-6.1) | 5.5(5.3-5.8) | 5.8(5.5-6.2) | 5.8(5.5-6.2) | 6.1(5.1-6.9) | <0.001 |
| U-P, g/gCr | | 1.14 (0.38-3.11) | 0.64 (0.14-2.82) | 1.09 (0.39-2.97) | 1.33 (0.59-2.74) | 4.99 (2.74-5.71) | 0.003 |
| Hematuria, % | | 289 (59.2%) | 42 (51.9%) | 174 64.4%) | 66 (51.6%) | 7 (77.8%) | 0.74 |
| Urine cast | | 266 (46.3%) | 24 (29.6%) | 129(47.8%) | 67 (52.3%) | 6 (66.7%) | 0.002 |
| Hypertension | | 227 (46.5%) | 19 (23.5%) | 119(44.1%) | 84 (65.6%) | 5 (55.6%) | <0.001 |
| DM | | 121 (24.8%) | 14 (17.3%) | 66 (24.4%) | 36 (28.1%) | 5 (55.6%) | 0.028 |
| ACE-I or ARB | | 171 (35.0%) | 10 (12.3%) | 101(37.4%) | 57 (44.5%) | 3 (33.3%) | <0.001 |
| Ca blocker | | 169 (34.6%) | 14 (17.3%) | 82 (30.4%) | 67 (52.3%) | 6 (66.7%) | <0.001 |
| Statins | | 116 (23.8%) | 14 (17.3%) | 68 (25.2%) | 31 (24.2%) | 3 (33.3%) | 0.39 |
| Smoking | | 211 (43.2%) | 26 (32.1%) | 119(44.1%) | 61 (47.7%) | 5 (55.6%) | 0.046 |
| Medullary fibrosis | 0 | 135 (27.7%) | 61 (75.3%) | 69 (25.6%) | 5 (3.9%) | 0 (0.0%) | <0.001 |
| | 1 | 239 (49.0%) | 18 (22.2%) | 160(59.3%) | 60 (46.9%) | 1 (11.1%) | |
| | 2 | 114 (23.4%) | 2 (2.5%) | 41 (15.2%) | 63 (49.2%) | 8 (88.9%) | |
| Medullary inflammatory cell infiltration | 0 | 359 (73.6%) | 78 (96.3%) | 217(80.4%) | 62 (48.4%) | 2 (22.2%) | <0.001 |
| | 1 | 107 (21.9%) | 3 (3.7%) | 52 (19.3%) | 50 (39.1%) | 2 (22.2%) | |
| | 2 | 22 (4.5%) | 0 (0.0%) | 1 (0.4%) | 16 (12.5%) | 5 (55.6%) | |
| Medullary cast formation | 0 | 254 (52.0%) | 74 (91.4%) | 152(56.3%) | 27 (21.1%) | 1 (11.1%) | <0.001 |
| | 1 | 170 (34.8%) | 7 (8.6%) | 93 (34.4%) | 68 (53.1%) | 2 (22.2%) | |
| | 2 | 64 (13.1%) | 0 (0.0%) | 25 (92.6%) | 33 (25.8%) | 6 (66.7%) | |

BMI, body mass index; sBP, systolic blood pressure; dBP, diastolic blood pressure; sCr, serum creatinine; eGFR, estimated glomerular filtration rate; HbA1c, hemoglobin A1c; U-P, urinary protein excretion; DM, diabetes mellitus; ACE-I, angiotensin–converting enzyme inhibitor; ARB, angiotensin II type 1 receptor blocker.

**Supplemental Table 5. Clinicopathologic diagnoses of the study population**

| Clinicopathologic diagnosis | n=488 |
|---|---|
| IgA nephropathy | 154 |
| ANCA-associated vasculitis | 49 |
| Lupus nephritis | 37 |
| MN | 35 |
| MCNS | 26 |
| Diabetic nephropathy | 26 |
| Nephrosclerosis | 24 |
| TIN | 23 |
| Minor glomerular changes | 19 |
| FSGS | 16 |
| IgA vasculitis | 16 |
| TBM | 14 |
| MPGN | 11 |
| Amyloid Nephropathy | 8 |
| Obesity-related nephropathy | 4 |
| TMA | 4 |
| IRGN | 3 |
| Cryoglobulinemic vasculitis | 3 |
| Alport syndrome | 3 |
| Immunotactoid nephritis | 2 |
| Fabry disease | 2 |
| Non-IgA mesangial proliferative glomerulonephritis | 1 |
| Anti-GBM glomerulonephritis | 1 |
| Light-chain proximal tubulopathy | 1 |
| Medullary cystic kidney disease | 1 |
| Polyarteritis nodosa | 1 |
| Lipoprotein glomerulopathy | 1 |
| BK virus–associated nephropathy | 1 |

ANCA, anti-neutrophil cytoplasmic antibody; MN, membranous nephropathy; MCNS, minimal change nephrotic syndrome; TIN, tubulointerstitial nephritis; FSGS, focal segmental glomerulosclerosis; TBM, thin basement membrane disease; MPGN, membranoproliferative glomerulonephritis; TMA, thrombotic microangiopathy; IRGN, infection-related glomerulonephritis; GBM, glomerular basement membrane. TIN includes sarcoid nephropathy and IgG4-related kidney disease. TMA includes calcineurin inhibitor-associated nephropathy.

**Supplemental Table 6. Spearman’s correlation coefficients among clinical and pathological variables**

| | Medullary fibrosis | | Medullary inflammatory cell infiltration | | Medullary cast formation | | IFTA | |
|---|---|---|---|---|---|---|---|---|
| | Correlation coefficient | p-value | Correlation coefficient | p-value | Correlation coefficient | p-value | Correlation coefficient | p-value |
| Age | 0.40 | <0.001 | 0.12 | 0.011 | 0.31 | <0.001 | 0.33 | <0.001 |
| BMI | 0.03 | 0.49 | 0.01 | 0.83 | -0.04 | 0.39 | -0.01 | 0.79 |
| sBP | 0.20 | <0.001 | 0.13 | 0.005 | 0.15 | <0.001 | 0.24 | 0.001 |
| dBP | 0.02 | 0.59 | 0.02 | 0.91 | 0.01 | 0.91 | 0.05 | 0.25 |
| eGFR | -0.53 | <0.001 | -0.38 | <0.001 | -0.48 | <0.001 | -0.57 | <0.001 |
| HbA1c | 0.17 | <0.001 | 0.04 | 0.44 | 0.09 | 0.055 | 0.16 | <0.001 |
| U-P | 0.11 | 0.017 | 0.07 | 0.13 | 0.14 | 0.002 | 0.13 | 0.004 |
| Medullary fibrosis | | | 0.46 | <0.001 | 0.54 | <0.001 | 0.56 | <0.001 |
| Medullary inflammatory cell infiltration | 0.46 | <0.001 | | | 0.30 | <0.001 | 0.42 | <0.001 |
| Medullary cast formation | 0.54 | <0.001 | 0.30 | <0.001 | | | 0.49 | <0.001 |
| IFTA | 0.56 | <0.001 | 0.42 | <0.001 | 0.49 | <0.001 | | |

BMI, body mass index; sBP, systolic blood pressure; dBP, diastolic blood pressure; eGFR, estimated glomerular filtration rate; HbA1c, hemoglobin A1c; U-P, urinary protein excretion; IFTA, interstitial fibrosis and tubular atrophy.

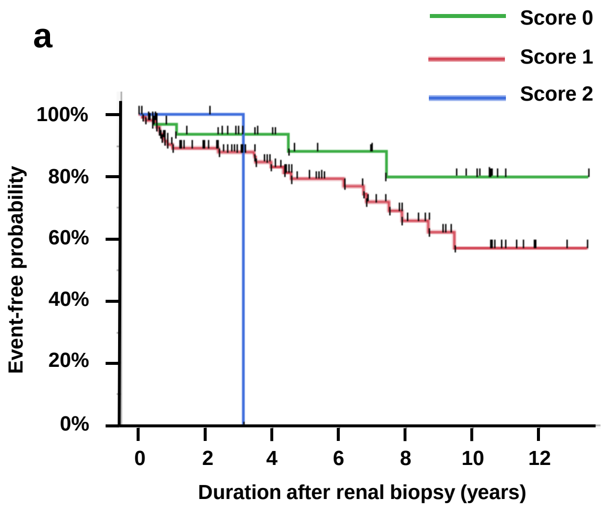


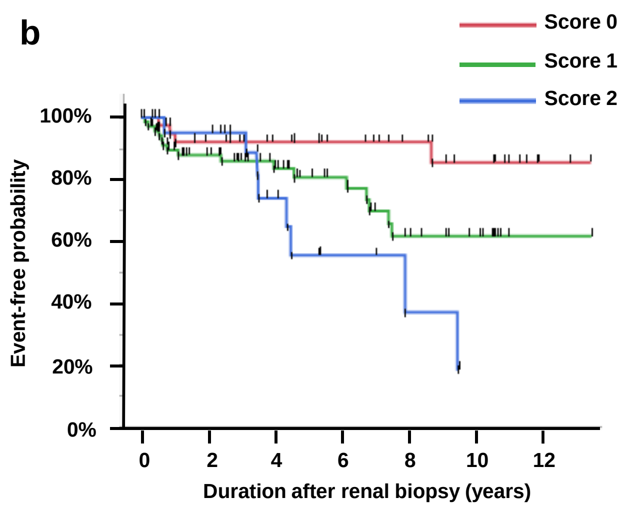


Number at risk

<Medullary inflammatory cell infiltration>

| | | | | | | |
|---|---|---|---|---|---|---|
| Score 0 | 113 | 71 | 49 | 33 | 21 | 11 |
| Score 1 | 34 | 27 | 19 | 14 | 10 | 8 |
| Score 2 | 2 | 2 | 0 | 0 | 0 | 0 |

Number at risk

<Medullary fibrosis>

| | | | | | | |
|---|---|---|---|---|---|---|
| Score0 | 53 | 32 | 25 | 20 | 15 | 10 |
| Score1 | 74 | 49 | 34 | 23 | 14 | 9 |
| Score2 | 22 | 19 | 9 | 4 | 2 | 0 |

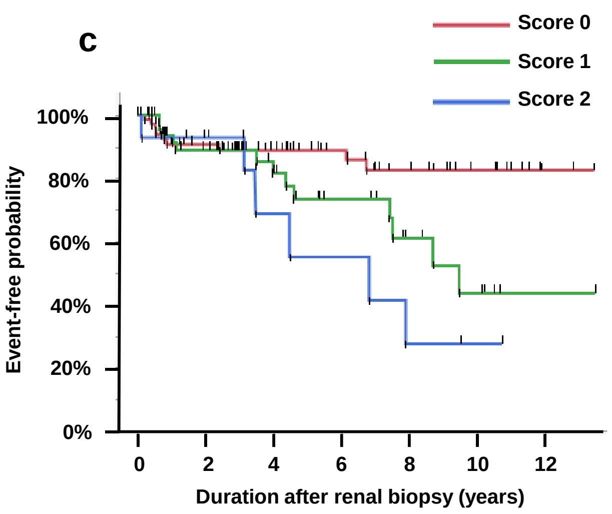


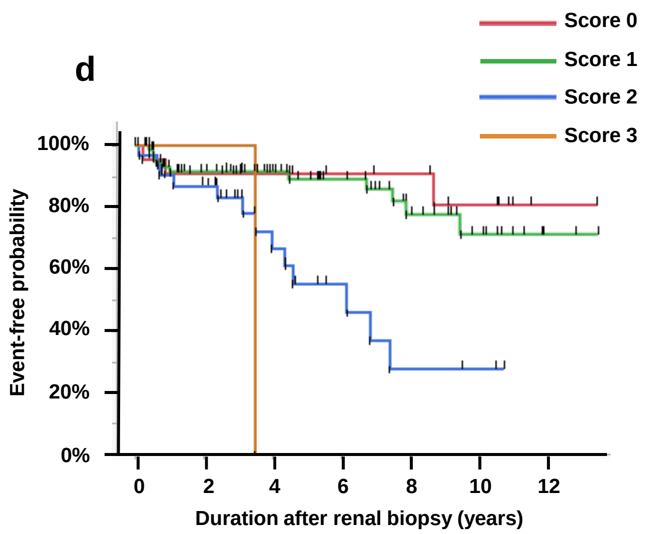


Number at risk

<Medullary cast formation>

| | | | | | | |
|---|---|---|---|---|---|---|
| Score 0 | 78 | 52 | 41 | 29 | 21 | 13 |
| Score 1 | 55 | 37 | 22 | 14 | 8 | 5 |
| Score 2 | 16 | 11 | 5 | 4 | 2 | 1 |

Number at risk

<IFTA>

| | | | | | | |
|---|---|---|---|---|---|---|
| Score 0 | 68 | 43 | 31 | 22 | 17 | 11 |
| Score 1 | 53 | 34 | 25 | 19 | 11 | 6 |
| Score 2 | 27 | 22 | 16 | 6 | 3 | 2 |
| Score 3 | 1 | 1 | 0 | 0 | 0 | 0 |

**Supplemental Figure 1. Renal survival rate in patients with IgA nephropathy.**

(a) Renal survival rate stratified by medullary inflammatory cell infiltration. Log rank test: p = 0.078. (b) Renal survival rate stratified by medullary fibrosis. Log rank test: p = 0.011. (c) Renal survival rate stratified by medullary cast formation. Log rank test: p = 0.016. (d) Renal survival rate stratified by IFTA score. Log rank test: p <0.001. IFTA, interstitial fibrosis and tubular atrophy.

**STROBE Statement**

| | Item No | Recommendation | Page No |
|---|---|---|---|
| **Title and abstract** | 1 | (*a*) Indicate the study's design with a commonly used term in the title or the abstract | 1-4 |
| | | (*b*) Provide in the abstract an informative and balanced summary of what was done and what was found | 3-4 |
| **Introduction** | | | |
| Background/rationale | 2 | Explain the scientific background and rationale for the investigation being reported | 5-6 |
| Objectives | 3 | State specific objectives, including any prespecified hypotheses | 6-7 |
| **Methods** | | | |
| Study design | 4 | Present key elements of study design early in the paper | 7 |
| Setting | 5 | Describe the setting, locations, and relevant dates, including periods of recruitment, exposure, follow-up, and data collection | 7 |
| Participants | 6 | (*a*) Give the eligibility criteria, and the sources and methods of selection of participants. Describe methods of follow-up | 7 |
| | | (*b*) For matched studies, give matching criteria and number of exposed and unexposed | 10 |
| Variables | 7 | Clearly define all outcomes, exposures, predictors, potential confounders, and effect modifiers. Give diagnostic criteria, if applicable | 9-10 |
| Data sources/ measurement | 8* | For each variable of interest, give sources of data and details of methods of assessment (measurement). Describe comparability of assessment methods if there is more than one group | 9 |
| Bias | 9 | Describe any efforts to address potential sources of bias | 9 |
| Study size | 10 | Explain how the study size was arrived at | 7 |

| | | | |
|---|---|---|---|
| Quantitative variables | 11 | Explain how quantitative variables were handled in the analyses. If applicable, describe which groupings were chosen and why | 10 |
| Statistical methods | 12 | (*a*) Describe all statistical methods, including those used to control for confounding | 10-11 |
| | | (*b*) Describe any methods used to examine subgroups and interactions | 13 |
| | | (*c*) Explain how missing data were addressed | N/A |
| | | (*d*) If applicable, explain how loss to follow-up was addressed | 10 |
| | | (*e*) Describe any sensitivity analyses | N/A |
| **Results** | | | |
| Participants | 13* | (a) Report numbers of individuals at each stage of study—eg numbers potentially eligible, examined for eligibility, confirmed eligible, included in the study, completing follow-up, and analysed | 11 |
| | | (b) Give reasons for non-participation at each stage | Figure 2 |
| | | (c) Consider use of a flow diagram | Figure 2 |
| Descriptive data | 14* | (a) Give characteristics of study participants (eg demographic, clinical, social) and information on exposures and potential confounders | Table 1, 12 |
| | | (b) Indicate number of participants with missing data for each variable of interest | |
| | | (c) Summarise follow-up time (eg, average and total amount) | |
| Outcome data | 15* | Report numbers of outcome events or summary measures over time | 12 |

| | | | |
|---|---|---|---|
| Main results | 16 | (*a*) Give unadjusted estimates and, if applicable, confounder-adjusted estimates and their precision (eg, 95% confidence interval). Make clear which confounders were adjusted for and why they were included | 12-13 |
| | | (*b*) Report category boundaries when continuous variables were categorized | 9 |
| | | (*c*) If relevant, consider translating estimates of relative risk into absolute risk for a meaningful time period | N/A |
| Other analyses | 17 | Report other analyses done—eg analyses of subgroups and interactions, and sensitivity analyses | 13 |
| **Discussion** | | | |
| Key results | 18 | Summarise key results with reference to study objectives | 15 |
| Limitations | 19 | Discuss limitations of the study, taking into account sources of potential bias or imprecision. Discuss both direction and magnitude of any potential bias | 17-18 |
| Interpretation | 20 | Give a cautious overall interpretation of results considering objectives, limitations, multiplicity of analyses, results from similar studies, and other relevant evidence | 15-17 |
| Generalisability | 21 | Discuss the generalisability (external validity) of the study results | 18 |
| **Other information** | | | |
| Funding | 22 | Give the source of funding and the role of the funders for the present study and, if applicable, for the original study on which the present article is based | 20 |
